# Lower than expected effective human-mosquito contact may regulate dengue virus transmission in high-suitability low-incidence environments: A plausible explanation for limited dengue outbreaks in Florida, U.S.

**DOI:** 10.64898/2025.12.09.25341886

**Authors:** Kristyna Rysava, Andrea Morrison, Gabriela Paz-Bailey, Danielle Stanek, Maile Thayer, Rebecca Zimler, Michael A. Johansson

## Abstract

Dengue, a vector-borne disease, puts at risk of infection nearly four billion people worldwide. Spread primarily by *Aedes* mosquitos, suitable mosquito habitat favoring survival, population maintenance, and an optimal extrinsic incubation period is required for successful transmission. Many areas, however, fall within the geographic range of the mosquito vector and favorable climatological conditions but experience only sporadic and short-lived outbreaks.

Here, we characterized local dengue virus transmission in Florida, United States, where transmission is limited despite frequent travel-associated cases and high climate-based estimates of the basic reproductive number. To reconcile the difference between expected R_0_ values and the observed transmission, we tested four transmission scenarios based on ecological assumptions on mosquito vector biology, human-mosquito contact, and temperature profiles. Specifically, we sought to understand the extent to which differences in effective contact between humans and mosquitoes may modulate the degree of expected transmission and assess variation between counties. Furthermore, we investigated possible drivers of differential contact rates between counties in the context of socioeconomic, demographic, and environmental variables.

We found that estimated human-mosquito contact rates across Florida were lower than those reported from endemic settings or estimated based on laboratory experiments, resulting in smaller outbreaks than expected given vector biology and local climatological conditions across the state. Associated explanatory variables moderating the degree of contact included population density and the built environment. The expected number of secondary cases generated using updated effective contact rates was in line with the existing epidemiological and genetic evidence, suggesting that low contact rates limit transmission from imported cases such that most result in no further transmission. These findings also imply that extended transmission risk is only present in southern Florida due to frequent travel-associated cases. The analytical framework presented here offers a new modifiable way to estimate the contact processes underlying dengue virus transmission in high-suitability low-incidence environments, which can be applied in other settings at the margins of endemicity, providing interpretable epidemiological insights instead of relying on traditionally used theoretical R_0_ values inferred from unmeasured entomological estimates.

## Introduction

Dengue, a vector-borne disease of major clinical significance, is a risk to nearly four billion people worldwide with an estimated 50-100 million cases of acute febrile illness every year (Cattarino et al., 2020; Brady et al. 2014). The causative agent of dengue fever, dengue virus (DENV), belongs to the *Flavivirus* genus along with Zika, yellow fever, and West Nile viruses, and exists in four antigenically distinct serotypes (Paz-Bailey et al., 2024; Reich et al., 2013). Spread by *Aedes aegypti* and *Ae. albopictus* mosquitos in tropical and sub-tropical regions, a suitable habitat favoring optimal population growth of the virus and its vector is required for DENV transmission. For example, precipitation, local hydrological conditions, and landscape character determine the quantity of pooled water bodies that provide the breeding sites for mosquitos to deposit their eggs (Lambrechts et al., 2011). In addition, the extrinsic incubation period, the time between when a mosquito takes an infected blood meal and when it becomes infectious and capable of spreading the virus onto a new host, varies with environmental temperature and is bound by the duration of the mosquito life cycle (Brady et al., 2013).

Whilst DENV transmission depends on the biological and environmental conditions, some areas which fall within the geographic range of *Ae. aegypti* and favorable climate experience only sporadic and short-lived outbreaks (Kraemer et al., 2015). Influx of infected humans combined with timing of suitable weather conditions are likely to play a critical role in determining if and to what extent DENV introductions will result in a substantial secondary transmission. In recent years, repeated DENV introductions, facilitated predominantly by travelers returning from dengue endemic countries, has led to localized emergence in many areas where the virus is not endemic. Within the contiguous United States (U.S.), the state of Florida has experienced the most frequent introductions and intense outbreaks of dengue and other arboviruses vectored by *Ae. aegypti* and *Ae. albopictus* such as Zika and chikungunya (CDC, 2023; Sharp et al., 2019; Bogoch et al., 2016; Kendrick et al., 2014).

Dengue cases are now routinely reported in individuals with recent travel history to dengue endemic countries. However, it was not until 2009 when Florida reported its first local outbreak since 1934. By the end of 2010, a total of 88 locally acquired infections were reported, followed by more outbreaks in 2013, 2020, and 2023 across several counties in southern Florida (Rowe et al., 2020; Teets et al., 2014; Graham et al., 2010). In addition, limited local transmission of Zika virus was documented in 2016 and 2017 as a result of the Zika epidemic across the Americas in 2015-2016 (Philip et al., 2019). Despite the steady importation of infected individuals and an increasing number of local transmission events, it remains unclear whether DENV circulates endemically at low prevalence or is limited to infrequent outbreaks, and what mechanisms may curtail local transmission.

*R*_0_, the number of secondary cases resulting from a single infection introduced into an entirely naïve population, is a common metric for infectious disease transmission risk and offers a helpful framework for assessing the likely intensity of a disease outbreak. For example, Mordecai et al. (2017) predicted an elevated risk of DENV transmission in Florida for ∼10 months a year based on the relationship between environmental temperature and mosquito biology. Similarly, by integrating information on the ecological niche of *Aedes* mosquitos and local temperature profiles, Bogoch et al. (2016) predicted transmission of Zika virus (ZIKV) in Florida, identifying the geographic region as conducive to autochthonous transmission all year round. Existing estimates of Flavivirus transmission, however, vary depending on whether the inference stems from biological and ecological underpinnings or directly from epidemiological data. In fact, by sequencing ZIKV genomes from infected patients and *Aedes aegypti* mosquitos and analyzing the actual reported cases, both Dinh et al. (2016) and Grubaugh et al. (2017) estimated low effective reproductive numbers (*R*_e_ of 0.16 and 0.5-0.8 respectively), suggesting that the locally acquired ZIKV infections were facilitated by multiple introductions rather than sustained local transmission, and that a decline of ZIKV was to be expected when imported cases decreased.

In theory, the differences between expected versus actual transmission (*R*_0_ and *R*_e_, respectively), primarily relate to immunity; however, that is unlikely to be the case in Florida. For immunity to have a significant impact on transmission, sustained local transmission would have to be common across many Florida counties reducing the pool of susceptible population. This type of transmission has not been detected in Florida, and studies of the age distribution of cases concur, estimating low transmission intensity and seroprevalence well below that of an endemic setting (Kada et al., 2024). From a climatological and ecological perspective, much of Florida is a highly suitable environment for DENV and the key *Aedes* vectors. The local epidemiology, however, appears limited to mostly sporadic cases or small outbreaks, with much less transmission than expected in other ecologically similar settings. Hence, the question remains of what mechanisms may drive the discrepancy between expected and actual transmission.

To investigate the potential gap in the theory, we first reviewed the equation traditionally used to calculate *R*_0_ of vector-borne diseases (Dietz, 1993):

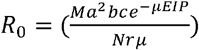

Here *M* and *N* refer to the mosquito and human population sizes, respectively. The parameter *a* indicates the mosquito human-feeding rate, *b* and *c* represent the probability of human-to-mosquito and mosquito-to-human infection given infectious contact, *μ* is the mosquito mortality rate, EIP is the extrinsic incubation period, and *r* is the duration for which humans are infectious. Most of the parameters related to mosquito vector biology are dependent on temperature and may differ by mosquito species. Moreover, all of the parameters can be difficult to ascertain, particularly from natural conditions as opposed to controlled experiments, and likely vary between settings. The parameter values are also typically characterized as averages, despite substantial heterogeneity across individuals that may contribute to transmission dynamics (Lloyd et al., 2007).

Given the significant temperature-dependent impact of the mosquito vector biology on DENV transmission, Mordecai et al. (2017) conducted an extensive literature review to estimate the compounded influence of temperature on *R*_0_ for *Ae. aegypti* and *Ae. albopictus* mosquito species. Whilst hugely informative for certain contexts, these estimations were based on an amalgamation of parameter values compiled from a set of laboratory observations and epidemiologically endemic backgrounds providing limited interpretability across diverse real-world settings. Moreover, the estimates of *R*_0_ were standardized to provide values between zero and one and used as a relative indicator of DENV transmission intensity rather than an actual basic reproductive number. As such, a further analysis of the *R*_0_ components is critical for understanding DENV transmission in Florida and similar, epidemiologically marginal or novel regions.

Specifically, mosquito abundance (*M*) is unknown and challenging to extrapolate across larger scales needed to estimate transmission risk across Florida. For that reason, many transmission models have also included mosquito population dynamics such as the population density model developed by Parham & Michael (2010) for which the probability of observing *M* mosquitoes at time *t* follows a Poisson distribution with mean equal to the ratio of temperature dependent birth and death rates of adult mosquitoes (new adults/lost adults). This approach, while attractive, requires assumptions such as uniform mosquito presence and population equilibrium, which do not capture the complex reality of spatiotemporal variation. In addition, the human population size (*N*) and disease recovery (*γ*) are often omitted from the final calculation, shifting the primary focus on vectoral capacity and disease reproductive potential, providing only biological interpretation of the transmission potential whilst ignoring the human side of an epidemiological interaction.

Similarly, the mosquito human-feeding rate is highly variable. This value is typically obtained from endemic settings or controlled experiments using diverse methodological approaches resulting in such estimates not only being less comparable amongst each other but also of limited use to novel settings such as Florida. Most studies of transmission risk have used assumptions based on either evidence of blood meals in trapped mosquitos (e.g., Lardeux et al., 2008 in Mordecai et al. 2017), human landing catch (Christofferson & Mores, 2011) or assuming that there is one blood meal per gonotrophic cycle (e.g., Frock & Barrera, 2002 in Mordecai et al. 2017). The abundance (or density) of adult female mosquitoes and the feeding rate together define a large part of the human-mosquito interaction that is critical for DENV transmission; yet both are variable, environmentally sensitive, and difficult to measure.

In this work we aimed to assess plausible mechanisms underlying effective human-mosquito transmission rate which may curtail DENV transmission risk in Florida. Instead of calculating the local transmission potential prospectively, we estimated biologically informed, time-specific, county-level basic reproductive numbers and their critical components by fitting a series of mechanistic models to over a decade of detailed surveillance data. We further explored the extent to which seasonal temperature variation and human-mosquito contact rates modulate this value. Given the existing body of literature, we hypothesized that the DENV transmission potential in Florida may be relatively high but substantially restricted by a combination of seasonal weather conditions and lower than expected contact between human and mosquito populations. The broader objective was to provide a quantitative framework to evaluate differences between individual settings similar in climatological and ecological conditions yet experiencing differential epidemiological outcomes regarding DENV transmission, to link these to potential socio-economic and behavioral drivers, and to identify knowledge gaps in vector transmission biology, particularly in non-endemic regions.

## Results

Between January 2009 and December 2022, Florida reported 303 locally acquired and 2,306 travel-associated cases. Most cases were located in southern counties with travel-associated cases exhibiting a wider geographical range (Figure 1). Specifically, 14% of counties (10/67) reported at least one locally acquired case and 62% of counties (46/67) reported at least one travel-associated case over the 14 years. In 2019 and 2022 Florida experienced unusually high numbers of travel-associated cases, in part associated with outbreaks in Cuba, leading to substantial secondary transmission particularly in Miami-Dade County (Embassy U.S., 2022; Sharp et al., 2019).

**Figure 1.**
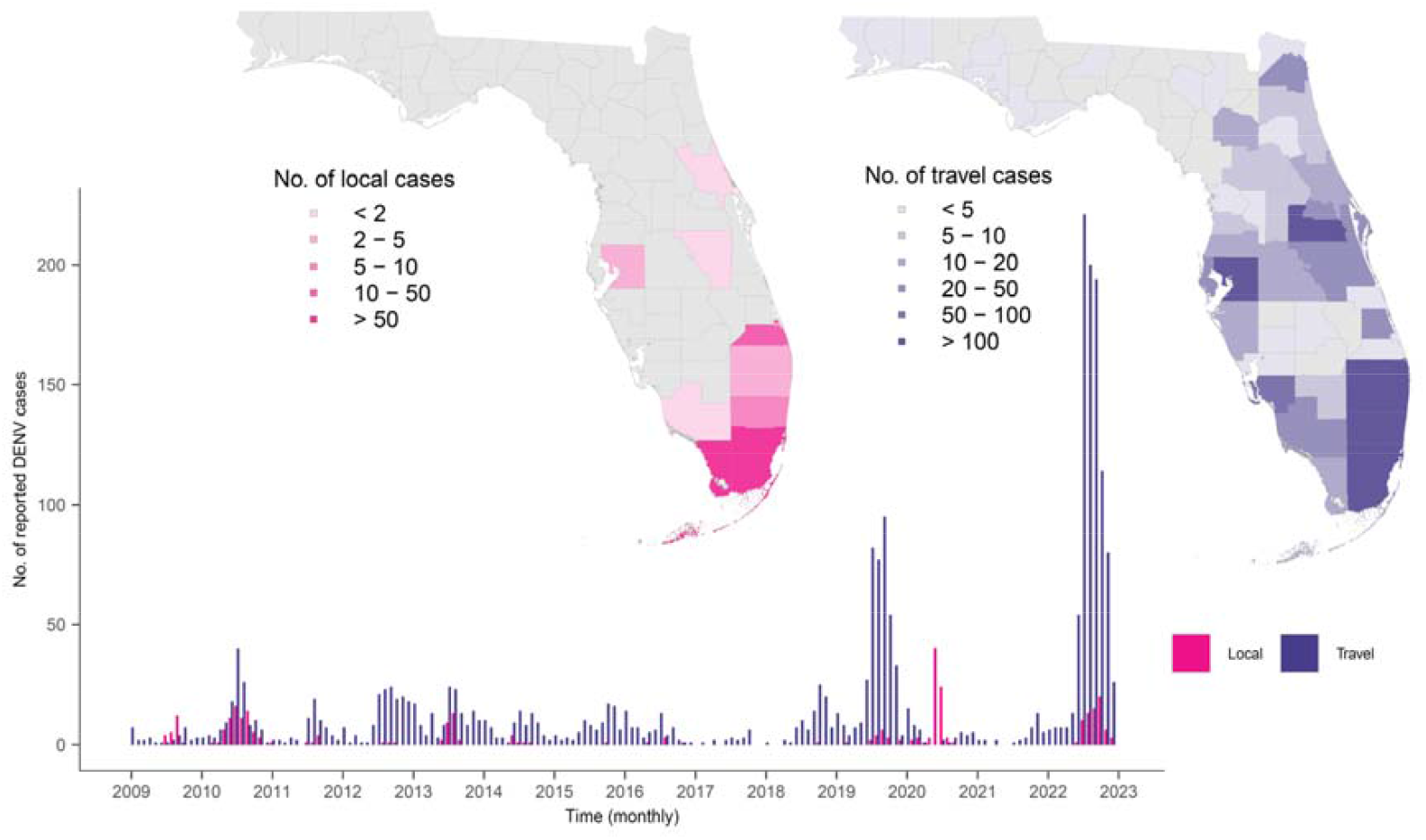
Geographical distribution and monthly time series of reported dengue cases in Florida from January 2009 through December 2022. Locally acquired cases are shown in magenta and travel-associated cases in purple. In the maps, counties are shaded by the total number of cases by local (left) and travel-associated (right) dengue exposure.

We developed a bi-weekly time step stochastic compartmental model with immune waning such that re-infection was possible after an average 6-month delay, mimicking possible susceptibility to subsequent infections by other DENV serotypes (Methods). We then explored the temporal trend in the locally acquired case trajectory by fitting variations of the model to the reported dengue case data from each county individually. We assessed four different transmission scenarios relative to the biology-informed archetypal vector transmission model (Introduction). In Scenario 1, we assumed that the effective human-mosquito contact rate (the combination of mosquito to human density ratio and the contact rate between humans and mosquitos), was unknown. In Scenario 2, we used a biologically informed biting rate and mosquito density based on an approach described in Johansson et al. (2014) and in the Methods section to pre-calculate time-specific *R*_0_ values following an updated formula in Mordecai et al. (2017). In addition, we allowed for the resulting transmission rate to change as transmission progresses, departing from a mass action transmission term by estimating an exponent to the mean number of new human infections (i.e., the rate of infection departs from the contact rate). This mechanism has been shown to better replicate epidemiological data for diseases like measles and rubella when discretizing the underlying continuous process, especially for mixing coefficients lower than unity due to spatial sub-structuring between the susceptible and infectious classes (Metcalf et al.; 2011; Bjornstad et al., 2002). Scenario 3 combined the previous two scenarios. In Scenario 4 we assumed the effective human-mosquito contact rate was unknown and that transmission had additional individual-level heterogeneity (i.e., a potentially long-tailed distribution).

All four models fit the dengue data reasonably well, predicting some local transmission every year in Miami-Dade County, despite several years having no reported cases (Figure 2). We found that Scenarios 1, 3, and 4 tended to estimate higher case numbers when outbreaks did occur and provided a better fit for Miami-Dade and most other counties, with respective total log-likelihood values across all fitted counties of −1448, −1453, and −1445 compared to −1536 for Scenario 2 (Table S1).

**Figure 2.**
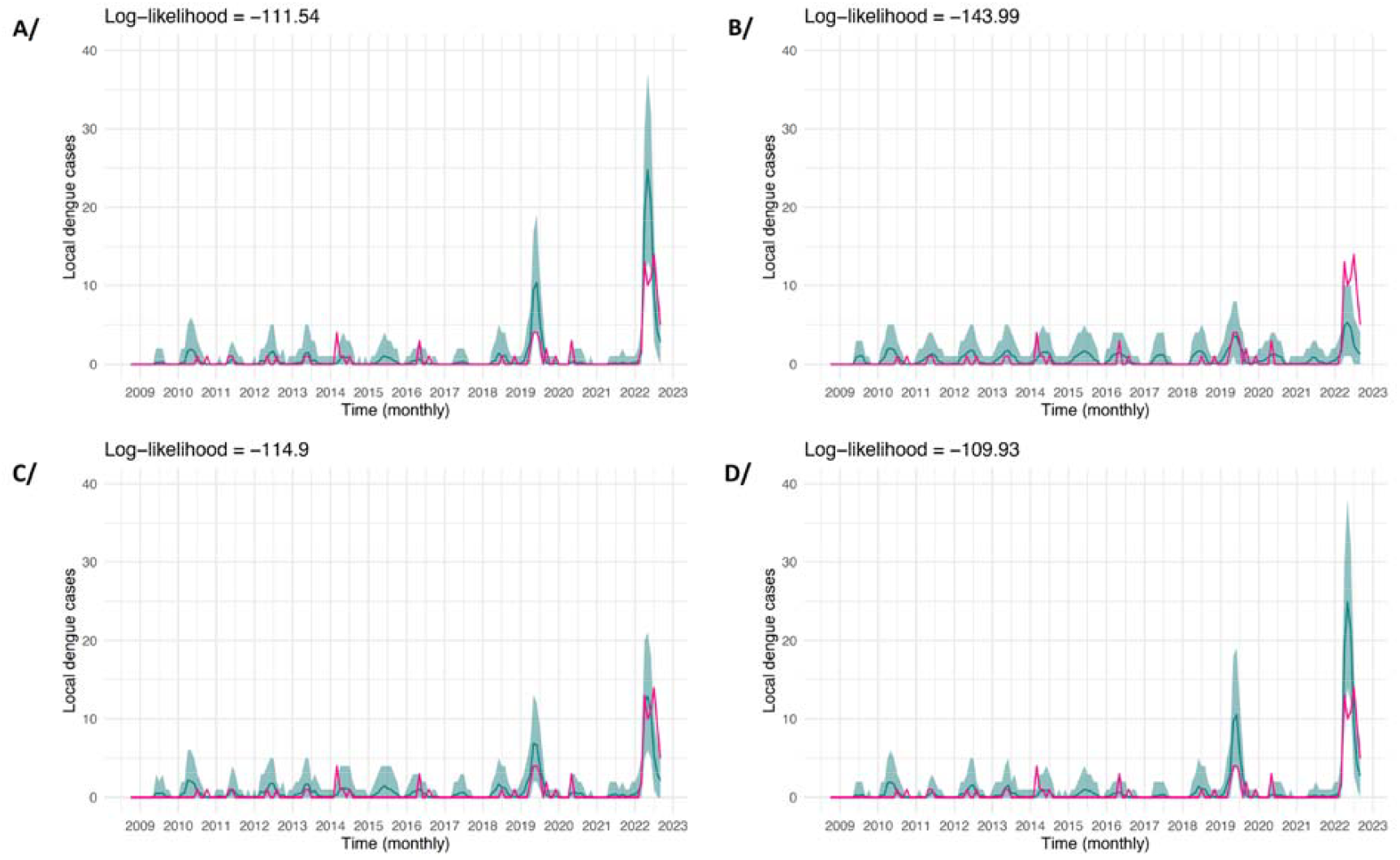
Resulting model fit for the four tested scenarios. Monthly time series figures of reported local dengue cases (magenta) in Miami-Dade County with fitted trajectories (turquoise, with the 95% prediction interval in lighter shading) from each dynamical model: **A/** Scenario 1: Effective contact unknown, **B/** Scenario 2: Biologically informed contact decreases as transmission progresses, **C/** Scenario 3: Effective contact unknown and decreases as transmission progresses, and **D/** Scenario 4: Effective contact unknown with additional individual-level heterogeneity.

The distributions of expected secondary infections varied across the scenarios, with a median of five cases expected in Scenario 2 and zero in Scenarios 1, 3, and 4 (Figure 3). Scenarios 1 and 4 were the most similar, with both a longer tail (up to 5 infections) and a higher proportion of no transmission events (approximately 90%). While actual transmission chains are difficult to observe, genetic and epidemiological evidence of transmission chains in Florida and particularly Miami-Dade County (Jones et al., 2024; Grubaugh et al., 2017) are in line with the pattern observed in Scenarios 1 and 4. For Scenario 1, the most parsimonious of these two scenarios, we estimated that 90% (95% quantile Q95 = 83 and 98%) of travel-associated cases would lead to no secondary case in Miami-Dade County, 5.3% (Q95= 1.24 and 8.17%) would result in a single secondary case, 3.1% (Q95=0.4. and 5.8%) would give rise to two secondary cases, 1.1% (Q95=0.1 and 2.3%) to three secondary cases, and 0.2% (Q95=0.0 and 0.7%) to four secondary cases. These findings are comparable with previous estimates based on temporal and phylogenetic relationships showing that 85% of travel-associated cases were potentially linked to zero locally acquired cases, 11% to a single case, and 1% being linked to 2-4 locally acquired cases during the DENV-3 outbreak in Florida between May 2022 and April 2023 (Jones et al., 2024).

**Figure 3.**
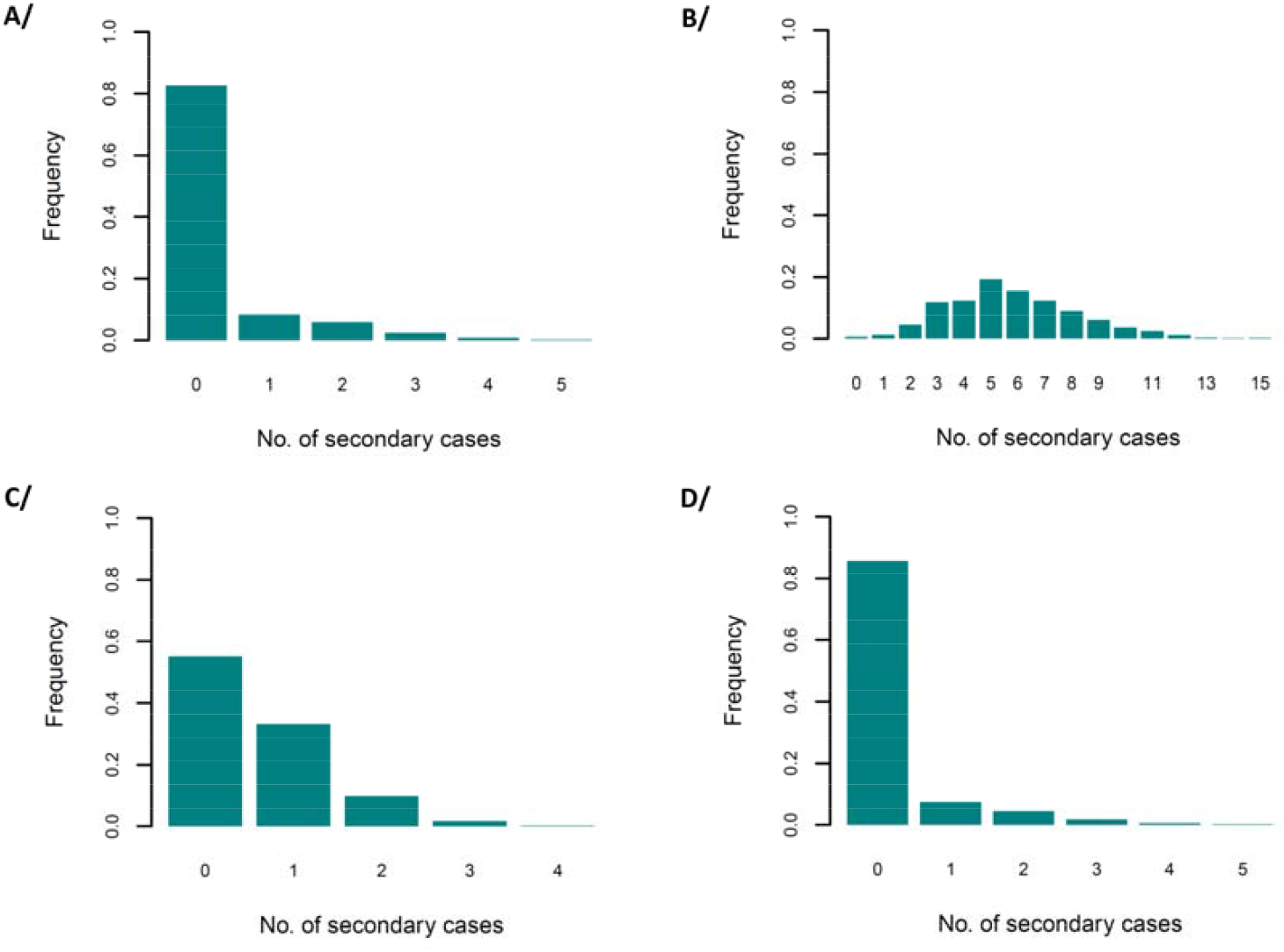
The expected number of locally acquired dengue cases following a single travel introduction in Miami-Dade County in summer for contact Scenarios 1-4 shown as **A/, B/, C/**, and **D/** in the same order.

Based on the parameter estimates from Scenario 1, we found that across counties and climatological seasons, the expected outbreak size and the number of generations per transmission chain following a single introduction were largely varied, with the highest transmission predicted for Monroe, Broward, and Miami-Dade counties in the summer season (Figure 4). The 95% quantiles from simulation studies showed a possibility of a larger outbreak in Monroe County specifically, illustrating a potential impact of higher effective human-mosquito contact. However, this risk was low and limited to summer and fall seasons, with most other counties showing minimal to no chance of sustaining an outbreak following a case importation due to minimal effective contact between humans and mosquitoes (Figure 5A). The average number of transmission generations in a chain was similarly limited, suggesting a stable relationship in the ratio between the outbreak size and the number of generations as opposed to a strong component of superspreading dynamics in the transmission of DENV in Florida (Figure 4B).

**Figure 4.**
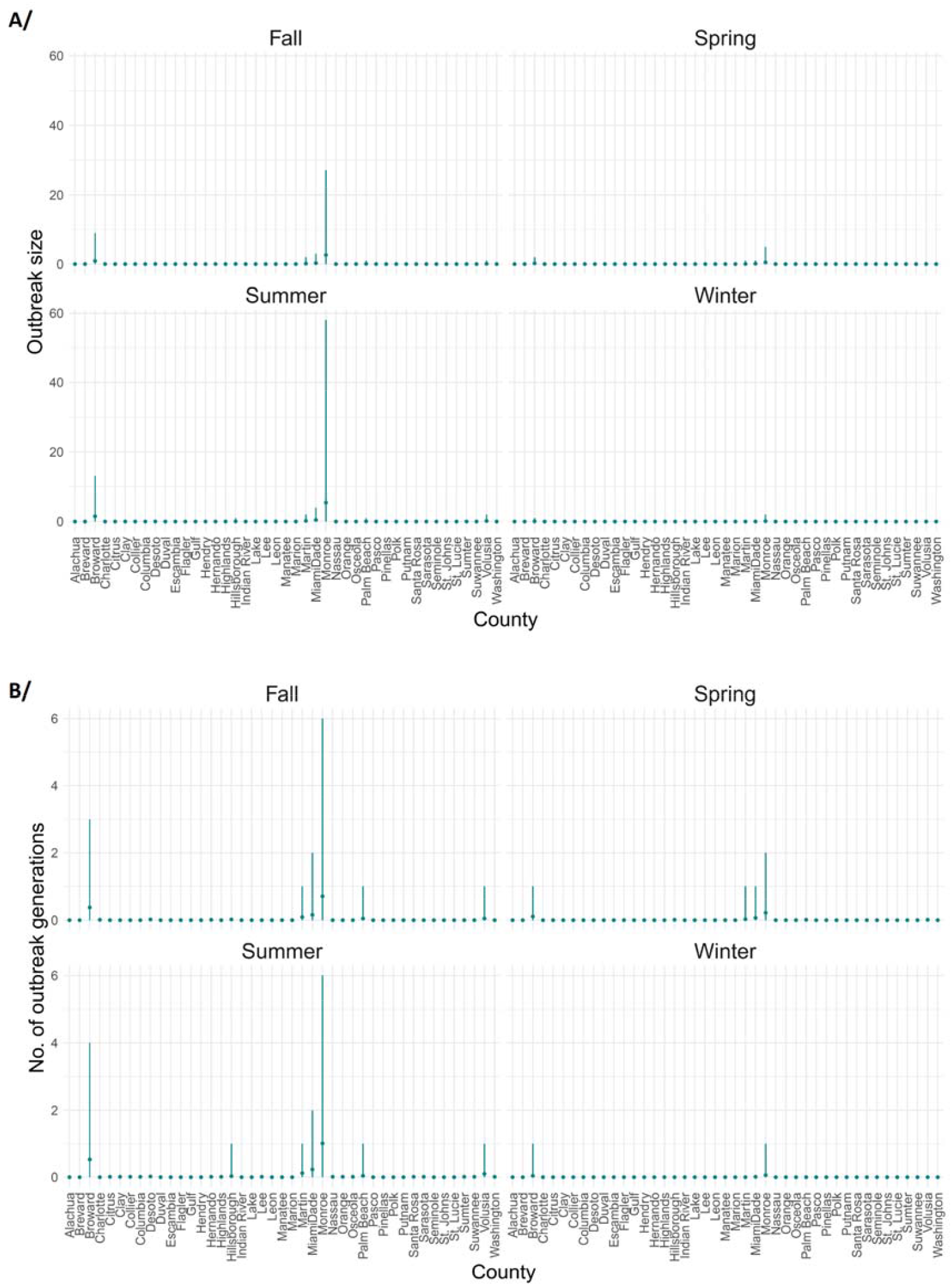
A summary of simulated transmission chains following a single travel introduction for Scenario 1 across Florida counties presented in the data and across climatological seasons. Mean **A/** outbreak size calculated as the number of all symptomatic and asymptomatic infections (assuming perfect observation) and **B/** outbreak length calculated as the number of generations of infections with 95% quantiles.

**Figure 5.**
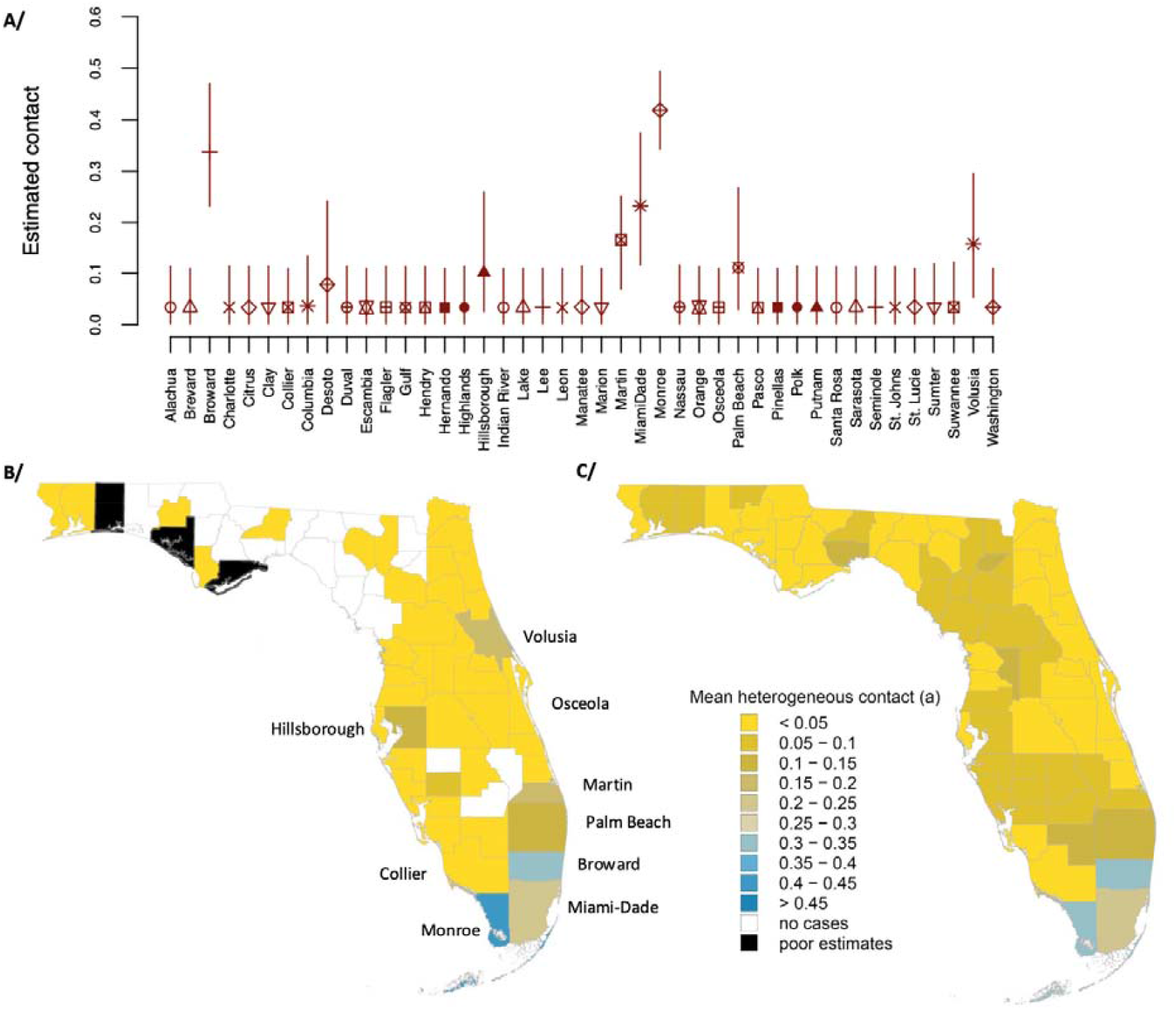
Estimated effective human-mosquito contact based on Scenario 1 across Florida counties. **A/** Effective contact rate by county with 95% confidence interval, **B/** choropleth of county effective contact rate estimated by the dynamical model, and **C/** effective contact rate estimated using the regression model with climatological and socio-economic explanatory variables (Table S2).

The estimated effective human-mosquito contact rate varied significantly across the counties, with higher rates concentrated in the southern tip of the peninsula (Figure 5b). We found that an increase in the county-level contact rates estimated using a weighted Gamma regression model was marginally associated with decreased population density (p-value < 0.01), decreased median number of rooms by tenure (p-value < 0.001), and increased proportion of occupied households (p-value < 0.01) A full list of regression variable p-values and model coefficients can be found in supplementary Table S2.

We further tested whether pre-existing immunity within the human population might modulate DENV transmission by re-running the dynamic model for Scenario 1 assuming 25% and 50% population-level immunity in Miami-Dade County, the county with the most travel-associated and locally acquired cases reported, and the highest percentage of foreign-borne population in Florida. We found that the log-likelihoods for all three models (0%, 25%, and 50% immunity) had comparable fits and estimated effective human-mosquito contact rates of 0.23 (0.11-0.37 for 95% quantile), 0.23 (0.13-0.43), and 0.32 (0.16-0.51) for the three tested levels of immunity (Figure S2). This indicates that the estimated contact rate had limited sensitivity to moderate levels of immunity in Miami-Dade and probably even less for other counties in which the levels of historical transmission or past exposure while travelling are likely significantly lower.

## Discussion

With changing mosquito ranges predicted for increased temperature scenarios, there is a potential for growing threat of DENV emergence in new settings. However, a susceptible human population, a competent mosquito vector, and frequent travel-associated introductions are already common across many areas of the Global North yet reported locally acquired dengue cases are limited despite general estimates of high dengue transmission potential in such areas. Therefore, the discrepancy between high suitability for and low incidence of the disease poses a question regarding potential mechanisms that are not captured in existing *R*_0_ estimates.

One key component of DENV transmission is the frequency of contact between unique humans and mosquitos, as a mosquito must acquire DENV from an infectious human and transmit it onward to a new susceptible human. Three approaches have been traditionally taken in existing modelling work to approximate the contact rate: (1) assuming a contact rate equal to the average number of blood meals taken by an infected mosquito per day, (2) equating the human landing catch with feeding, or (3) assuming a contact rate equivalent to one contact per mosquito gonotrophic cycle reflecting the biological requirement of at least one blood meal for reproduction. However, our understanding of these biological mechanisms is limited due to challenges in observing such behaviours in the field and across different ecological and epidemiological settings. Moreover, they are intrinsically different. If mosquitoes feed multiple times per blood meal, which has been shown for *Ae. aegypti* (Scott et al., 2000), then the gonotrophic cycle is an underestimate of the frequency of blood meals. On the other hand, if a mosquito feeds exclusively on one person, there may be no risk of transmission regardless of how many meals they take. As such, even a perfect understanding of the mosquito biology would not necessarily translate to transmission risk without detailed empirical contact-tracing data.

Similarly, estimates of mosquito abundance in the field are limited and hard to extrapolate across larger spatial aggregates given highly differential land-use at fine scales. Therefore, in most existing models the number of exposed humans per infected mosquito is either derived from theory as described in Parham & Michael (2010) (see Introduction) or from simulation models (e.g., Focks et al., 1993). Such approximations, however, take on other significant biological assumptions regarding spatial and temporal homogeneity and completely omit the role of human behavior, mosquito host preference, and the built environment such as human-made spaces and urban infrastructures. Alternative approaches focus on estimating the absolute disease reproductive potential of an adult mosquito in the absence of information on population sizes (Obolski et al., 2019). Such estimates are informative of the amplitude and timing of transmission potential but limited in their interpretability regarding epidemic growth and, when confronted with data, they often fail to explain transmission patterns driven by a wider range of epidemiological components (e.g., case importations). As such, most existing models used to predict dengue emergence either employ parameters capturing mosquito vector biology based on smaller-scale studies in endemic settings and controlled experiments or miss important contextual factors altogether.

Here, we explicitly estimated human-mosquito contact patterns for a non-endemic area to understand dengue dynamics in high-suitability low-prevalence settings. Specifically, we employed case data from the state of Florida, U.S., leveraging its spatiotemporally heterogenous case incidence and detailed travel surveillance documenting frequent DENV introductions. By testing a series of transmission scenarios, we found that despite the relatively high theoretical risk of DENV transmission, actual transmission was likely limited by a combination of seasonality and lower than expected effective contact rates between humans and mosquitoes. Whilst the effective contact rate presented here, and the biting rates reported in previous studies are not directly comparable, their epidemiological consequences are. In fact, we demonstrated that lower effective contacts limit DENV transmission in Florida despite suitable climate – an outcome in concord with the reported case incidence but at odds with the previously predicted high DENV risk. We further narrowed down the set of plausible biological processes constraining the degree of human-mosquito disease interaction below those assumed in traditional climate-driven models. Consequently, our modelling framework unifies the very mechanisms that are likely to influence transmission that would be otherwise logistically challenging to obtain at scale and resolution, making such an epidemiological inference possible.

Potential socio-ecological mechanisms reducing the intensity of effective mosquito-human contact include variations in human-mosquito mixing possibly due to use of air-conditioning or window screens, minimal amount of time spent outside, variable levels of mosquito control, and mosquito biting host preference and availability. Previous evidence suggested that lifestyle may limit DENV transmission by shifting mosquito host feeding patterns to non-human species (Olson et al., 2020). For example, lower density housing in Texas was associated with lower DENV incidence, possibly interrupting feeding between households (Reiter et al., 2003). Similarly, Olsen et al. (2021) showed that locations with lower human population densities were more likely to experience “aegyptism without arbovirus” – a phenomenon describing *Ae. aegypti* environmental suitability with no endemic DENV transmission. In the meta-analysis of the dynamical model outcomes here, we found a different trend, that counties with lower population density tended to have higher estimated effective human-mosquito contacts. This finding together with higher estimated contact rates in counties with fewer rooms per household and higher occupancy may reflect a complementary mixture of socioeconomic factors influencing risk in higher density locations in Florida. For example, these factors could reflect densely populated counties with higher frequency of high-rise and apartment buildings, seasonal residences, and limited outdoors space which could reduce mosquito-human contact. This study is not designed or powered to provide further insight on what could drive these differences. Further investigations into the relationship between the human-mosquito infectious contact, population density, cultural and lifestyle habits, and the housing and landscape management are needed for more conclusive answers.

Furthermore, heterogeneity in human-mosquito mixing may play an additional role in curtailing population-level DENV transmission (Perkins et al., 2013). Whilst here we used a larger-scale model that does not directly assess specific mechanisms of heterogeneity, it likely does capture some of the same components in the average human-mosquito contact rate and, in contrast to more complex models, can be fit to large-scale epidemiological data. Notably, Scenario 4, which included additional transmission event level heterogeneity, did not fit the data any better than Scenario 1, in which we estimated only effective contact rates. Scenario 3, which uses a model originally trialed to address heterogeneity in measles virus transmission fit notably worse than the other models as the changes in observed dynamics indicate reduced transmission with higher number of infections rather than at the point of single introductions.

In fact, DENV transmission in Florida appears to be principally driven by travel-associated cases. Many counties received one or more travel-associated cases over the course of 14 years, but for continued local transmission, a sustained supply of imported cases was generally required. Therefore, the intrinsic risk of transmission was only relevant for a limited number of counties in southern Florida where temperature, precipitation, and our estimates of effective human-mosquito contact were relatively high. Monroe and Martin counties, both of which appear to have had only small local outbreaks, are among the counties with higher estimated effective contact rates with limited secondary transmission predicted to occur in the summer months when transmission is expected to be most successful. Another possible factor limiting DENV transmission is pre-existing dengue immunity in counties with substantial reports of travel-associated cases. Whilst the role of immunity was only assessed in a sensitivity analysis here, the levels of immunity estimated in Monroe County (5.4% tested positive for recent dengue infection in CDC, 2010) and Florida (estimated seroprevalence for 35-year-olds of 9.6% in Kada et al., 2024), were unlikely to limit transmission in Florida at scale. However, it is possible that in particular contexts such as seasonal travel destinations or certain sub-populations, varying degrees of immunity may impact localized transmission intensity and susceptibility.

Broadly speaking, the low incidence of dengue in Florida and other southern states across the U.S. may be primarily due to socioeconomic and behavioral factors despite the suitable climate and mosquito populations. However, it is the socioeconomic context shaping the degree of human-mosquito contact that warns against generalized conclusions. Whilst in many places around the world living conditions and daily habits may evolve in response to global change, such as increases in air-conditioned environments to escape increasingly hotter temperatures, the impact of shifts in temperature with respect to disease risk remain highly uncertain and varied. Populations and sub-populations with lower socioeconomic privilege may not have the same opportunities to adapt and may therefore experience increased dengue exposure and subsequent disease (Farinelli et al., 2018). Whilst this is a public health concern in its own right, it also stresses the importance of functioning surveillance and control systems given that endemic foci of infection may act as a source of introductions increasing the global likelihood of infection across the areas with suitable climate and mosquito populations.

The analyses conducted here identify a new potential mechanism for restricted DENV transmission in Florida. By integrating setting-specific, travel-associated and locally acquired case data with mechanistic models without relying on assumed mosquito-human parameters, we offer a novel modifiable way to estimate the contact processes underlying DENV transmission in high-suitability low-incidence environments. Our modelling framework can be applied in other settings at the margins of endemicity, further identifying differences in contact between human and mosquito populations under a range of spatiotemporal and ecological contexts, providing interpretable epidemiological insights instead of depending on theoretical R_0_ values inferred from unmeasured entomological estimates. Targeted field studies collecting site-specific information on human-mosquito contact obtained through a standardized sampling approach and accompanied by potential explanatory variables (such as mosquito species and host abundance, built environment factors, and host behavioral patterns) would be an ideal addition to understand how the human-mosquito contact is modified. Such studies would lead to improvements on current models and, inherently limited model assumptions. Beyond shedding more light on the mechanisms of DENV transmission in non-endemic settings they could also help disentangle the spatial scale at which these modulators operate, ultimately leading to better mitigation strategies to disease management.

## Methods

I. **Data** Dengue case data were provided by the Florida Department of Health that conducts both passive and active case-finding activities for DENV in response to reported cases, with IgM and reverse transcription PCR testing for case confirmation as applicable. Florida Department of Health routinely interviews suspected case-patients for possible risk factors (e.g. travel, mosquito bite exposure) and possible mosquito exposure locations (for locally acquired cases), as well as attempts to identify additional symptomatic contacts preceding or succeeding the interviewed case (Rowe et al., 2020). The data provided were, however, spatially resolved by county level only for personal security reasons. We further aggregated the reported case data temporarily by Julian bi-weeks from January 2009 to December 2022, differentiating between locally acquired and travel-associated cases given the most likely infection origin. Whilst syndromic surveillance intensified during 2022 with focus on travel activities from Cuba, for most of the study period, there were no major changes in surveillance or diagnostic practices, and we assumed reporting probabilities were consistent over time. To collect climatological data such as temperature and precipitation for each Florida county we utilized the ‘EpiNOAA’ package in the programming environment R and aggregated these observations by bi-weeks to match the temporal resolution of the epidemiological dataset. Information on the county or household level sociodemographic variables was then procured through the ‘tidycensus’ package from 2009 to 2022 and averaged over the time period of the study, each as a potential driver of the by county human-mosquito effective contact rate.
II. **Temperature-sensitive dengue transmission model & regression analysis** To estimate the degree of local DENV transmission in Florida, we developed a stochastic, single-host discrete time, compartmental model expanded by several critical features informed from the biology and ecology of the pathogen and its host populations. Here we applied basic demographics with human individuals leaving the population through natural mortality *µ* and new susceptible individuals to be born at the rate, *b*. Human infection induced immunity waned after an average of 6 months after which recovered individuals migrated back into the susceptible compartment. Travel-associated introductions were drawn from a negative binomial distribution with size equal to reported travel cases *C*_*Tt*_ and reporting probability *ρ*_*T*_ as

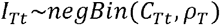

The process model time step *t* was modeled bi-weekly, while locally acquired cases were observed at four-week intervals (to reduce noise in the bi-weekly data) with a binomial reporting probability as

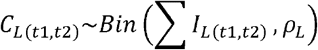

Here *C*_*L*_ indicate the reported local cases given all infected local cases *I*_*L*_ in the system were observed with reporting probability *ρ*_*L*_ during the last two bi-weeks (*t*1, *t*2). We fitted all mechanistic models to data using maximum likelihood calculated via iterated filtering algorithm for estimating parameters of partially observed Markov processes using an R package ‘pomp’ (Euler method for numerically solving stochastically in King, Nguyen, and Ionides 2015) in the programming environment R. For all scenarios, the parameters fitted were travel and local reporting probabilities. For Scenarios 1,3 and 4 we also fitted the effective human-mosquito infectious contact, for Scenarios 2 and 3 the exponent of mass action departure, and for Scenario 2 only the mosquito biting rate.
  A. For Scenario 1, we focused on estimating the county-level effective human-mosquito infectious contact rate *ε* as

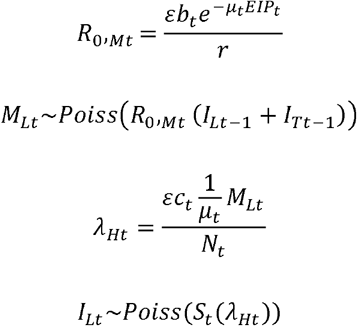

Here *R*_0,*Mt*_ and *λ*_*Mt*_ generally indicate transmission potential for human and mosquito populations, with *M*_*Lt*_ and *I*_*Lt*_ indicating local infectious mosquitoes and humans at the time *t* (bi-week), respectively. Specifically, *λ*_*Ht*_ is calculated as 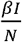, *where* 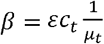 *and I = M*_*LT*_, whilst *R*_0, *Mt*_, represents the value of an average number of secondary infections caused by a single infectious mosquito defined as *R*_0_ *= βγ where* 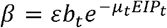 *and* 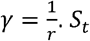 and *N*_*t*_ denote susceptible and total human populations at the same time step as described above. The effective contact rate *ε* is a free parameter.
  B. In Scenario 2, we assumed the overall transmission potential to be curtailed as transmission progresses due to localized density dependence as

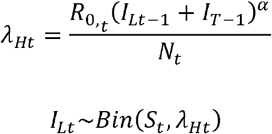

Here, *α* captures potential localized depletion of susceptible contact at the global scale by limiting transmission at elevated levels of predicted incidence across the county-level population. Human population parameters remained as described above. 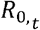 in this instance equals *β* as 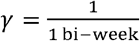, and was calculated following an adapted formula and most functional parameters for *Ae. aegypti* taken from Mordecai at al., 2017:

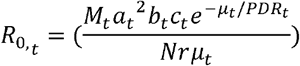

Note that PDR here refers to the parasite development rate which is the inverse of the extrinsic incubation period (EIP). The per-mosquito biting rate *a* as well as the human to mosquito ratio (M/N) were taken from Johansson et al., 2014. Specifically, the maximized ratio under ideal temperature conditions was assumed to be two mosquitos per person with increased mosquito mortality at temperature extremes such as

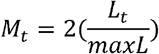

Where *L*_*t*_ denotes the temperature-dependent mosquito lifespan and *maxL* is the mosquito maximum mean lifespan of 7.9 days (Johansson et al., 2014; Brady et al., 2013).
  C. For Scenario 3, we combined elements of the previous two mechanisms (Scenarios 1 and 2) and estimated the effective infectious contact *ε* together with the dampening coefficient *α* as

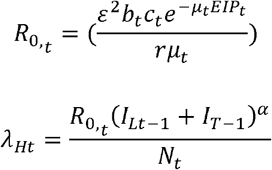

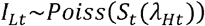

Here again, *β* and 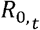 take on the same value as 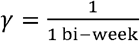.
  D. In Scenario 4, we further expanded Scenario 1 by allowing additional flexibility in the form of a dispersion parameter *k* when generating the number of new infectious human and mosquito individuals from negative binomial distribution as

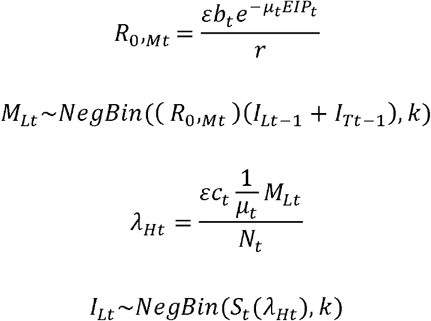

In all four scenarios parameters *b*_*t*_ and *c*_*t*_ indicate the probability of human-to-mosquito and mosquito-to-human infection given infectious contact, and parameters *EIP*_*t*_ and *µ*_*t*_ refer to the mosquito extrinsic incubation period and mosquito mortality respectively. All, except for mosquito mortality for which we followed a formula provided in Johansson et al., 2014, were pre-calculated following the thermal response model in Mordecai et al., 2017, given bi-weekly average temperatures obtained for each county. Human recovery rate *γ* was assumed to be an inverse of an average of 5 days infectious period, *r* (WHO, 2024).

Whilst a pre-existing immunity is unlikely to play a critical role in the dynamics of DENV transmission in Florida given the average disease incidence, we conducted an additional sensitivity analysis. As most locally acquired and travel-associated cases are reported in Miami-Dade County, we tested whether an improvement in the model fit would be achieved by including immunity levels of 25% and 50% within the county. We assumed no improvement in the model fit under increased immunity levels in Miami-Dade to indicate the lack of immunity’s importance across Florida.

Lastly, we examined potential drivers of differential infectious contact between counties by fitting the estimated contact rate from the best-fit scenario (Scenario 1) to Gamma regression linear model weighted by the number of travel-associated introductions with information on county-level temperature and precipitation, household management, socioeconomic, and immigration background and primary mosquito species as possible explanatory variables. The full model after checking for variable collinearity by calculating variance inflation factors (VIF<5) included the following terms: minimum temperature, minimum precipitation, human population density, median household income, per capita income, median number of rooms per household, proportion of households occupied, proportion of households owned, proportion of U.S. citizens born abroad of American parent(s), proportion of non-U.S. citizens, and county-level primary mosquito species. For final model selection, we used a stepwise backward model selection algorithm based on model AIC values utilizing the “stepAIC” function in R package ‘MASS’. All analyses were performed in the programming environment R version 4.4.0 (R Core Team, 2021).

## Data Availability

Code and data available on GitHub.

https://github.com/IstyRsquared/Dengue-Contact.git

