## Supplementary material for "Lower than expected effective human-mosquito contact may regulate dengue virus transmission in high-suitability low-incidence environments: A plausible explanation for limited dengue outbreaks in Florida, U.S."

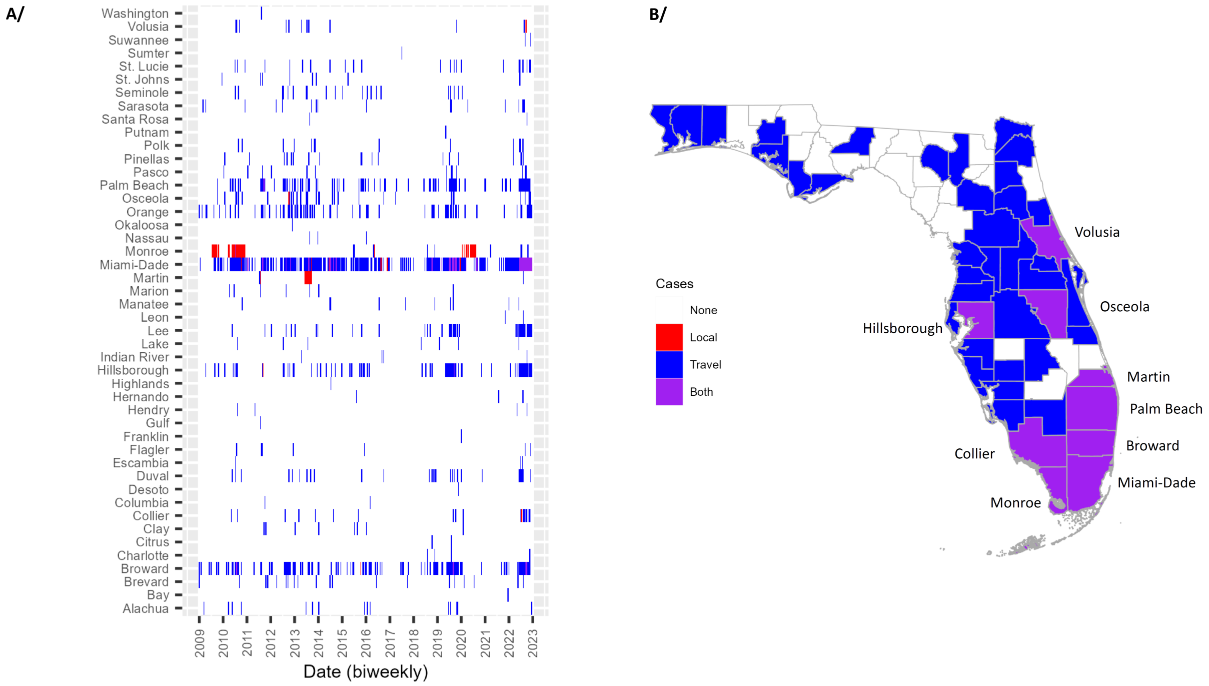


**Figure S1.** Dengue cases in Florida between January 2009 and December 2022. **A/** Bi-weekly case presence by county. Locally acquired cases are indicated by red, travel-associated cases by blue, and the occurrence of both case types in a given 2-week period shown in purple. Whilst many counties experienced repeated introductions, only very few reported locally acquired secondary transmission. **B/** Spatial distribution of Florida counties with reported dengue cases. Blue polygons show counties which received dengue introductions from outside but did not report any local transmission, while purple polygons point to areas with both case types reported.

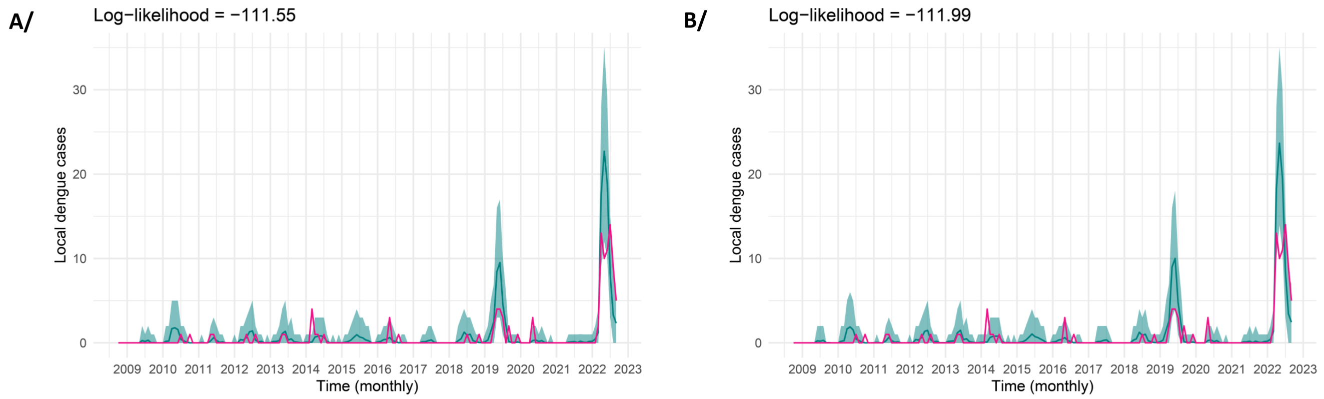


**Figure S2.** Monthly time series figures of local dengue cases in Miami-Dade County with fitted trajectories from the dynamical model for Scenario 1 in which we estimated human-to-mosquito contact across Florida counties. Here **A/** and **B/** demonstrate the output of sensitivity analyses assuming 25% and 50% dengue immunity in the county's population, respectively, with the loglikelihood values shown at top of each plot.

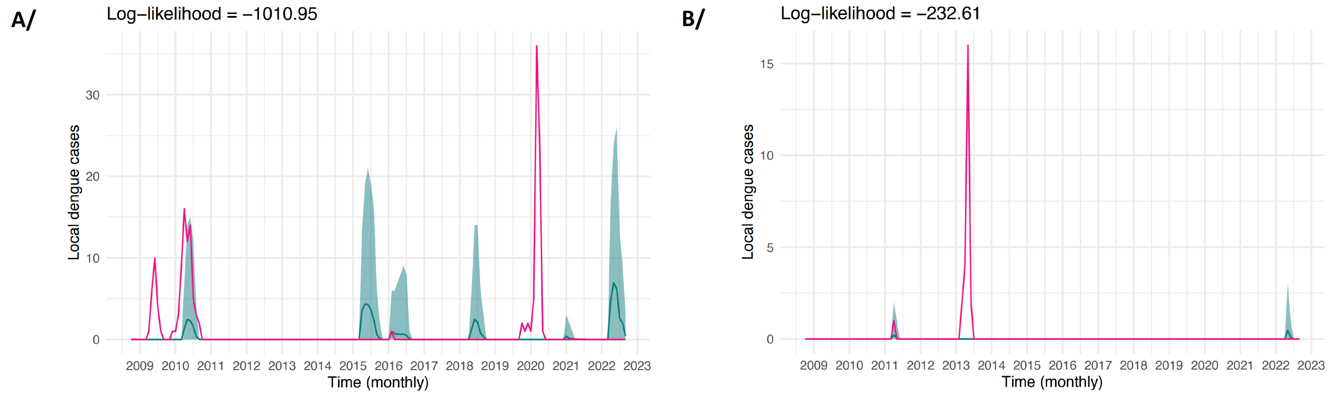


**Figure S3.** Monthly time series figures of local dengue cases in **A/** Monroe and **B/** Martin counties. For both local outbreaks, the 2020 outbreak in Monroe and 2013 outbreak in Martin, there were no travel-associated cases reported giving to the model's local incidence underestimates around the time.

**Table S1.** Loglikelihood values of dynamical model fit for Scenarios 1-4 across Florida counties.

| County | Scenario 1 | Scenario 2 | Scenario 3 | Scenario 4 |
|---|---|---|---|---|
| Alachua | 0 | -0.96 | 0 | 0 |
| Bay | 0 | 0 | 0 | 0 |
| Brevard | 0 | -2.13 | 0 | 0 |
| Broward | -17.96 | -18.25 | -16.91 | -17.74 |
| Charlotte | 0 | -0.47 | 0 | 0 |
| Citrus | 0 | -0.23 | 0 | 0 |
| Clay | 0 | -0.33 | 0 | 0 |
| Collier | 0 | -1.91 | 0 | 0 |
| Columbia | 0 | -0.01 | 0 | 0 |
| Desoto | 0 | 0 | 0 | 0 |
| Duval | 0 | -1.5 | 0 | 0 |
| Escambia | 0 | -0.49 | 0 | 0 |
| Flagler | 0 | -0.47 | 0 | 0 |
| Franklin | 0 | 0 | 0 | 0 |
| Gulf | 0 | -0.18 | 0 | 0 |
| Hendry | 0 | -0.99 | 0 | 0 |
| Hernando | 0 | -0.68 | 0 | 0 |
| Highlands | 0 | -0.27 | 0 | 0 |
| Hillsborough | -7.19 | -8.72 | -6.83 | -7.17 |
| Indian River | 0 | -0.34 | 0 | 0 |
| Lake | 0 | -1.15 | 0 | 0 |
| Lee | 0 | -3.18 | 0 | 0 |
| Leon | 0 | -0.1 | 0 | 0 |
| Manatee | 0 | -1.38 | 0 | 0 |
| Marion | 0 | -0.94 | 0 | 0 |
| Martin | -232.61 | -233.4 | -232.42 | -232.63 |
| MiamiDade | -111.54 | -143.99 | -114.9 | -109.93 |
| Monroe | -1010.95 | -1023.48 | -1013.88 | -1010.64 |
| Nassau | 0 | -0.11 | 0 | 0 |
| Okaloosa | 0 | 0 | 0 | 0 |
| Orange | 0 | -4.99 | 0 | 0 |
| Osceola | -57.56 | -59.56 | -57.56 | -57.56 |
| Palm Beach | -7.19 | -10.09 | -7.5 | -7.21 |
| Pasco | 0 | -1.11 | 0 | 0 |
| Pinellas | 0 | -1.9 | 0 | 0 |
| Polk | 0 | -1.42 | 0 | 0 |
| Putnam | 0 | -0.35 | 0 | 0 |
| Santa Rosa | 0 | -0.12 | 0 | 0 |
| Sarasota | 0 | -1.55 | 0 | 0 |
| Seminole | 0 | -2.26 | 0 | 0 |
| St. Johns | 0 | -0.63 | 0 | 0 |
| St. Lucie | 0 | -2.21 | 0 | 0 |
| Sumter | 0 | -0.26 | 0 | 0 |
| Suwannee | 0 | -0.03 | 0 | 0 |
| Volusia | -2.62 | -3.63 | -2.64 | -2.59 |
| Washington | 0 | -0.13 | 0 | 0 |
| Total | -1447.62 | -1535.88 | -1452.65 | -1445.48 |

**Table S2.** Weighted Gamma regression model summary statistics. The final model null deviance equals 1149.153 on 42 degrees of freedom and residual deviance of 92.091 on 30 degrees of freedom.

| Variable name | Exponentiated coefficient | Coefficient | p-value | Predicted contact with 1-unit increase in variable |
|---|---|---|---|---|
| Intercept | 0.003 | -5.774 | 0.357 | 0.003 |
| Mean temperature | 1.337 | 0.291 | 0.087 | 0.004 |
| Mean precipitation | 2.012 | 0.699 | 0.26 | 0.006 |
| County human population density | 0.298 | -1.211 | 0.006 | 0.001 |
| Median household income | 1 | 0 | 0.142 | 0.003 |
| Median number of rooms per household | 0.067 | -2.709 | 0 | 0 |
| Per capita income | 1 | 0 | 0.052 | 0.003 |
| Proportion of households occupied | 509.39 | 6.233 | 0.003 | 1.582 |
| Proportion of households owned | 21.937 | 3.088 | 0.266 | 0.068 |
| Proportion of U.S. citizens born abroad to American parents | 2.46E+38 | 88.398 | 0.242 | 7.64E+35 |
| Proportion of non-U.S. born residents | 0.002 | -6.183 | 0.056 | 0 |
| Primary mosquito species Ae. albopictus | 0.49 | -0.714 | 0.098 | 0.002 |
| Primary mosquito species mixed Ae. aegypti & albopictus | 1.005 | 0.005 | 0.973 | 0.003 |
